# Changes in Depressive Symptom Domains During Treatment with Accelerated and Conventional Repetitive Transcranial Magnetic Stimulation

**DOI:** 10.64898/2026.08.03.26359627

**Authors:** Michael R. Apostol, Thomas E. Valles, Juliana Corlier, Michael K. Leuchter, Alexander S. Young, Hewa Artin, Ralph J. Koek, Evan H. Einstein, Scott A. Wilke, Hanadi A. Oughli, Thomas Strouse, Aaron Slan, Margaret G. Distler, Dustin Z. DeYoung, Nathaniel Ginder, David E. Krantz, Andrew F. Leuchter

## Abstract

Accelerated 5x5 repetitive Transcranial Magnetic Stimulation (rTMS; five stimulation sessions per day for five days) is an effective treatment for Major Depressive Disorder (MDD), and has efficacy comparable to conventional once-daily rTMS. Considering the heterogeneity of symptoms in patients with MDD, it is critical to determine how accelerated 5x5 and conventional rTMS affect depression symptom domains. We compared symptom change over time in patients treated with either accelerated 5x5 rTMS (25 total sessions, *n* = 40) or conventional once-daily rTMS administered over six weeks (30 total sessions, *n* = 135). Accelerated 5x5 patients received either prolonged intermittent theta burst stimulation (piTBS) or personalized “resonant frequency” (RF) stimulation. Mixed-effects linear models were built to compare the two protocols, with the primary outcome variables being the Inventory of Depression Symptomology Self-Report (IDS), the Ruminative Response Scale (RRS), and the Profile of Mood States - Brief (POMS), yielding measures of 14 unique depression symptom domains. Both protocols led to similar improvements in all 14 depression symptom domains (all interaction term *p*-values > .05). Subsequent exploratory analyses demonstrated that accelerated 5x5 and conventional rTMS may differ in the time courses of their effects on anxiety, rumination, mood, depression, and vigor (*p*-values < .05, uncorrected). These results suggested that accelerated 5x5 rTMS has a similar efficacy in alleviating 14 depression symptom domains compared to conventional once-daily rTMS, and that either protocol may be appropriate for MDD patients with a variety of symptom profiles.

## Introduction

Major Depressive Disorder (MDD) is the leading cause of disability worldwide (Abdoli et al., 2022; Belmaker & Agam, 2008; Cui et al., 2024; Otte et al., 2016; World Health Organization, 2017). Patients with MDD display variability in onset, progression, severity, and symptom presentation (aan het Rot et al., 2009; Chen & Saito, 2021; Chu et al., 2023; Lohoff, 2010; Lynch et al., 2020; Marx et al., 2023; Schatzberg et al., 2000; Wang et al., 2012). The heterogeneity of depression symptoms between patients poses a significant challenge for stratifying patients to effective interventions (Cuijpers et al., 2020; Tozzi et al., 2024; Williams, 2024).

MDD research often defines the severity of depression through overall sum scores (Fried & Nesse, 2015a), an approach that may not capture the heterogeneity of depression (Bahr et al., 2008; Fried, 2017; Fried & Nesse, 2015b). MDD patients with comparable overall severity may have significantly different individual symptom profiles (Lynall & McIntosh, 2023) and analysis of separate depression symptoms may capture symptom heterogeneity between patients (Williams et al., 2024). For example, anger is present in MDD patients and is related to increased disease severity and duration (Judd et al., 2013). It is possible that a more tailored approach to treatment selection could help to target particular symptom clusters more effectively (Williams & Whitfield Gabrieli., 2025).

Repetitive Transcranial Magnetic Stimulation (rTMS) is effective in patients with treatment-resistant depression (M. T. Berlim et al., 2013; M. Berlim et al., 2013; Dell’Osso et al., 2011; Fitzgerald & Daskalakis, 2013, 2022; Klomjai et al., 2015), often targeting the left dorsolateral prefrontal cortex (L-DLPFC) (Blumberger et al., 2018; Downar & Daskalakis, 2013; Hsu et al., 2024; Perera et al., 2016; Caulfield & Brown, 2022; Moretti & Rodger, 2022). Two contrasting approaches to rTMS treatment are “conventional” (once per day, five days per week) and “accelerated” (multiple treatments per day over several consecutive days) protocols. Accelerated approaches may help patients achieve remission from MDD in a shorter period of time (Lefaucheur et al., 2025; Cole et al., 2020, 2022). Conventional and accelerated rTMS protocols have similar tolerability, safety, and efficacy (Caulfield et al., 2022) for overall depression treatment outcomes (Brunoni et al., 2017; Cole et al., 2022; Kaster et al., 2019), but it is unclear whether conventional and accelerated rTMS have distinct effects on heterogeneous depression symptoms between patients (Kaster et al., 2023). Indeed, recent work has demonstrated that different depression symptom domains show varying time courses of improvement throughout rTMS treatment (Einstein et al., 2026). Determination of whether conventional and accelerated rTMS protocols affect different depression symptom domains differently may help improve treatment personalization (Chen & Saito, 2021; Chu et al., 2023; Lynch et al., 2020; Marx et al., 2023; Schatzberg et al., 2000; Thaipisuttikul et al., 2014).

We recently reported that conventional once-daily rTMS had similar efficacy to accelerated 5x5 rTMS (5 sessions per day for 5 consecutive days) in reducing overall depression scores, with 5x5 rTMS producing responses in less than 1 week (Apostol et al., 2026). Here, we extend this work by comparing the effects of conventional and accelerated 5x5 rTMS on different depression symptom domains that are common in MDD patients (Liang et al., 2025; Kaźmierczak et al., 2026; Marx et al., 2023). We examined the effects of conventional versus accelerated rTMS protocols on changes in mood, anxiety, sleep, anhedonia, life engagement, rumination, depression, anger, confusion, fatigue, tension, and vigor. We hypothesized that although both protocols reduced overall symptom severity (Apostol et al., 2026) they may have differential effects on specific depression symptom measures.

## Methods

### Patient Population

Data were collected from 175 patients with MDD who received rTMS at the UCLA TMS Clinical and Research Service (Los Angeles, CA, USA). Patients who received rTMS from February 2023 to March 2025 were included in this study. The Mini International Neuropsychiatric Interview (Sheehan et al., 1998) was used to confirm a diagnosis of MDD. 135 patients underwent conventional rTMS, which included one treatment per day, five days a week, for 6 weeks (30 rTMS treatments total; Einstein et al., 2026; M. K. Leuchter et al., 2023). 40 patients received accelerated 5x5 rTMS which consisted of either piTBS (*n* = 25; Cole et al., 2020, 2022; Slan et al., 2024) or individualized resonant frequency (RF) rTMS (*n* = 15; A. F. Leuchter et al., 2021). Accelerated 5x5 rTMS was administered five times per day for five consecutive days (25 sessions total). This study was approved by the UCLA Institutional Review Board. Data were collected in accordance with the Declaration of Helsinki (World Health Organization, 2013). Informed consent was obtained from patients who received RF-rTMS as part of a separate ongoing study.

### Clinical Assessments

Patients completed self-report clinical assessments prior to the initiation of rTMS, every 5 rTMS sessions (i.e., once per week for conventional patients and once per day for accelerated patients), and again after the completion of the last rTMS session. Assessments were completed by participants using a tablet device. Patients completed the Inventory of Depression Symptomology Self-Report (IDS; Rush et al., 1996; Trivedi et al., 2004), which measured overall depression symptoms and associated symptoms including mood, anxiety, sleep (Wardenaar et al., 2010), life engagement (Thase et al., 2023), and anhedonia (the current study included a modified IDS Anhedonia subscale based on Fukuda et al., 2021). The IDS overall scores ranged from 0 (minimal depression) to 84 (severe depression). The Ruminative Response Scale (RRS; Townshend & Hajhashemi, 2023) was collected as a measurement of depressive rumination with scores ranging from 22 to 88, where higher scores reflect increased rumination. Finally, patients completed the Profile of Mood States – Brief (POMS; Petrowski et al., 2021), which is a measure of various moods including tension, depression, anger, fatigue, confusion, and vigor. Each POMS subscale has a maximum score of 20. See Table 1 for the specific measure items included in each subscale.

**Table 1.**
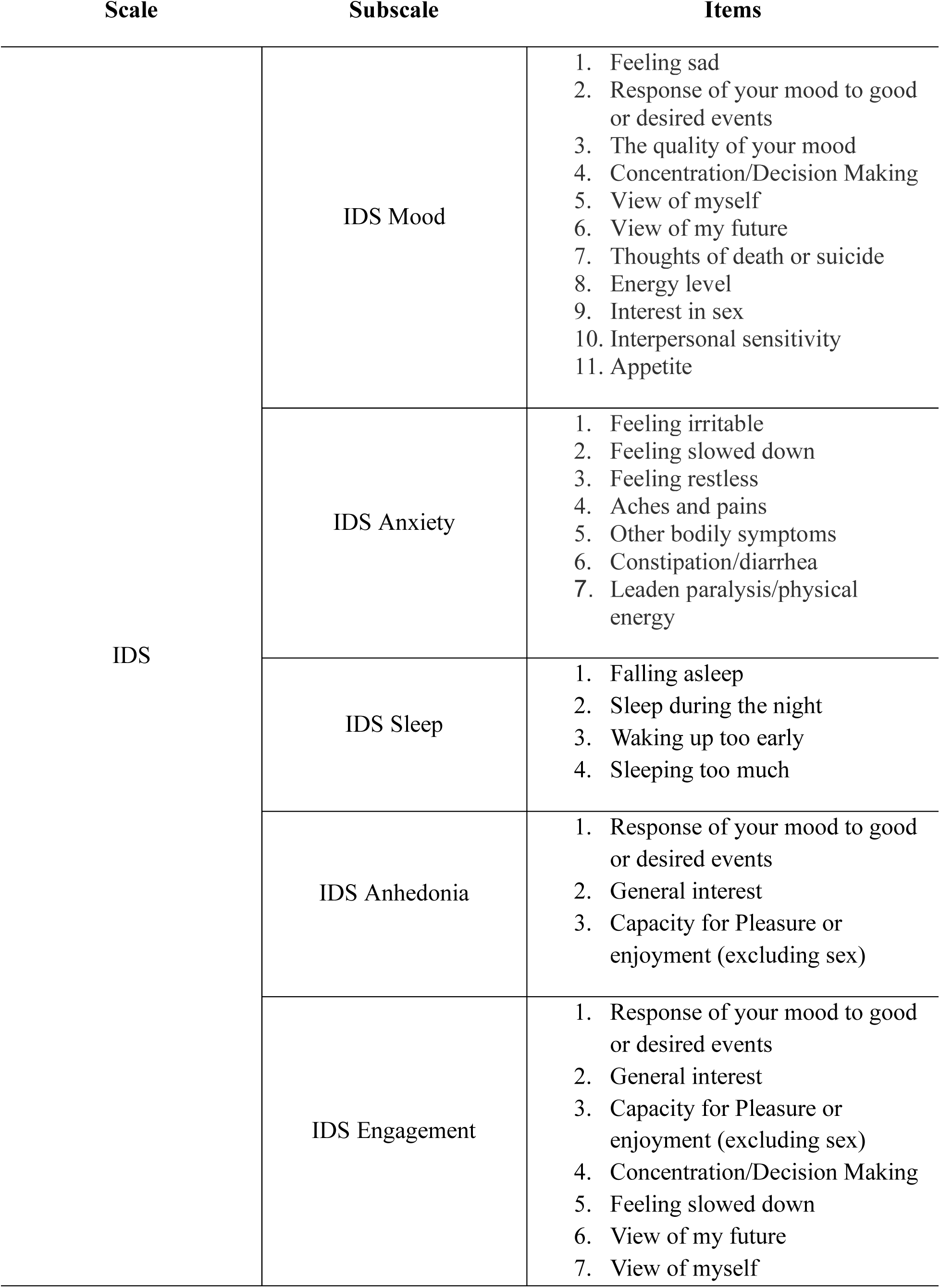
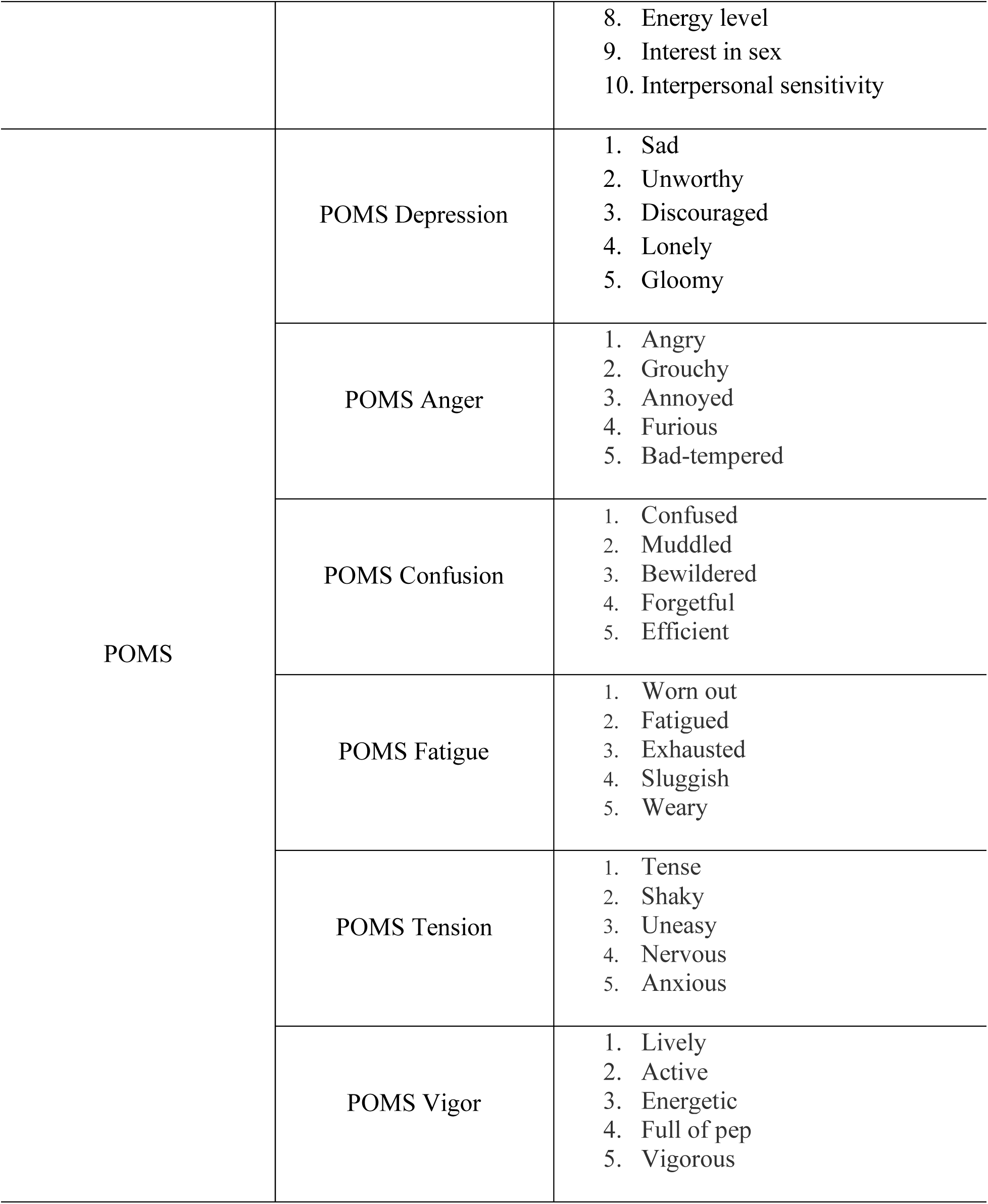
The questionnaire items that were used to generate each subscale.

### rTMS Procedures

Patients received rTMS from one of three rTMS devices: the Magstim Super Rapid2 (Magstim Inc., MN, USA), Magstim Horizon (Magstim Inc., MN, USA), or MagPro X100 (MagVenture, Inc., GA, USA) outfitted with a figure-eight rTMS coil. Each patient underwent a resting motor threshold (rMT) procedure which was defined as the maximum stimulator intensity that elicited a motor evoked potential in the right abductor pollicis brevis muscle for at least 5 of 10 single rTMS pulses applied to the contralateral primary motor cortex.

Conventional rTMS included stimulation once per day, five days per week, for six weeks (30 total sessions). Of the 135 patients who received conventional rTMS, *n* = 132 began their treatment course with 3000 pulses of 10 Hz rTMS and *n* = 3 began their course with 1800 pulses of piTBS. The stimulation intensity was increased to 120% as tolerated from treatments one to five. The stimulation target was the L-DLPFC which was localized with the Beam F3 method (Beam et al., 2009). A measurement-based care paradigm was used to adjust stimulation parameters in patients who displayed limited improvement in depression symptom severity or who were unable to tolerate stimulation (Apostol et al., 2026; Chu et al., 2023; Citrenbaum et al., 2023; Fitzgerald et al., 2009; Lee et al., 2020; M. K. Leuchter et al., 2023). For example, patients may have received bilateral DLPFC stimulation or completed procedures to increase treatment tolerability.

Accelerated 5x5 rTMS included five sessions per day for five consecutive days (25 total sessions). Patients received piTBS based on clinician referral (*n* = 25) or RF-rTMS by consenting to enroll in an ongoing study focused on RF-rTMS (*n* = 15). piTBS treatment included a 9-minute stimulation session of 1800 pulses. RF-rTMS patients underwent a “frequency mapping” procedure using electroencephalography (EEG) measures to identify the stimulation frequency that was associated with the greatest predicted improvement in MDD symptoms (A. F. Leuchter et al., 2021). RF-rTMS patients received 3000 pulses delivered in 40-pulse trains at their individualized RF, which ranged from 6-17 Hz. Accelerated 5x5 treatments had an inter-session-interval of 45 minutes. During the waiting period between treatments, patients rested in a private waiting room and could engage in quiet activities. As with conventional rTMS, the stimulation intensity was increased to 120% as tolerated from treatments one to five.

## Data Analysis

Analyses were performed using R version 4.4.1 in RStudio (Allaire et al., 2012). Pre-treatment characteristics were compared between the conventional and accelerated patients using t-tests with age, IDS, RRS, and POMS as outcome variables.

To test whether there were differences in the changes in depression subscale scores after conventional versus accelerated rTMS, mixed-effect linear models were built for the following outcome variables: IDS overall score, a series of IDS subscales (anhedonia, anxiety, mood, sleep, and engagement), RRS, POMS overall score, and a series of POMS subscales (confusion, depression, fatigue, tension, and vigor). It is important to note that increases in the POMS vigor subscale is a desirable outcome, whereas decreases in all the remaining subscales is a sign of improvement in depression symptom domains following rTMS. Each outcome variable was included in a separate model with timepoint (pre versus post rTMS) and treatment type (accelerated 5x5 versus conventional rTMS) as fixed effects. Participants were included as a random effect. Mixed-effects linear models were built using the *lmerTest* package in R (Kuznetsova et al., 2017). “Post-rTMS” was defined as measures taken after session 30 for the conventional patients and after session 25 for the accelerated patients, reflecting a complete treatment course (Apostol et al., 2026). For these primary analyses, the Bonferroni correction (.05 / 14 = .0036) was applied.

Exploratory models were then built to examine longitudinal changes in depression symptom domain measures during the rTMS treatment course. Mixed-effect linear models were built with the same outcome variables, fixed effects, and random effects as the primary analyses, with one exception: the “timepoint” fixed effect now included all time points where measures were taken (baseline, after sessions 5, 10, 15, 20, and 25). The last timepoint included was treatment 25 because this was the final measure both conventional and accelerated patients completed. The exploratory analyses were designed to identify if differences arise between conventional and accelerated 5x5 rTMS prior to the completion of a full treatment course (e.g., how might IDS sleep subscale scores differ between conventional and accelerated patients after treatment 10?). Exploratory analyses were not corrected for multiple comparisons and are viewed as hypothesis-generating. As such, exploratory models should be interpreted conservatively.

## Results

The conventional treatment (average age 46.69. female 71, male 62, non-binary 2; baseline IDS = 41.35) and accelerated treatment patients (average age 49.90, female 22, male 18’ baseline IDS = 41.30) did not differ statistically in baseline age or depression symptom scores (age: *p* = 0.316; IDS: *p* = 0.981; RRS: *p* = 0.660; POMS: *p* = 0.6389. This suggests that the patients who received accelerated 5x5 versus conventional rTMS were comparable in terms of age and depression symptom severity prior to treatment. Trajectories of symptom change demonstrated that both rTMS treatment types led to improvements in the IDS, IDS subscales, RRS, POMS, and POMS subscales, but that improvements in the accelerated patients occurred in five days, compared to six weeks for the conventional patients (Figure 1).

**Figure 1.**
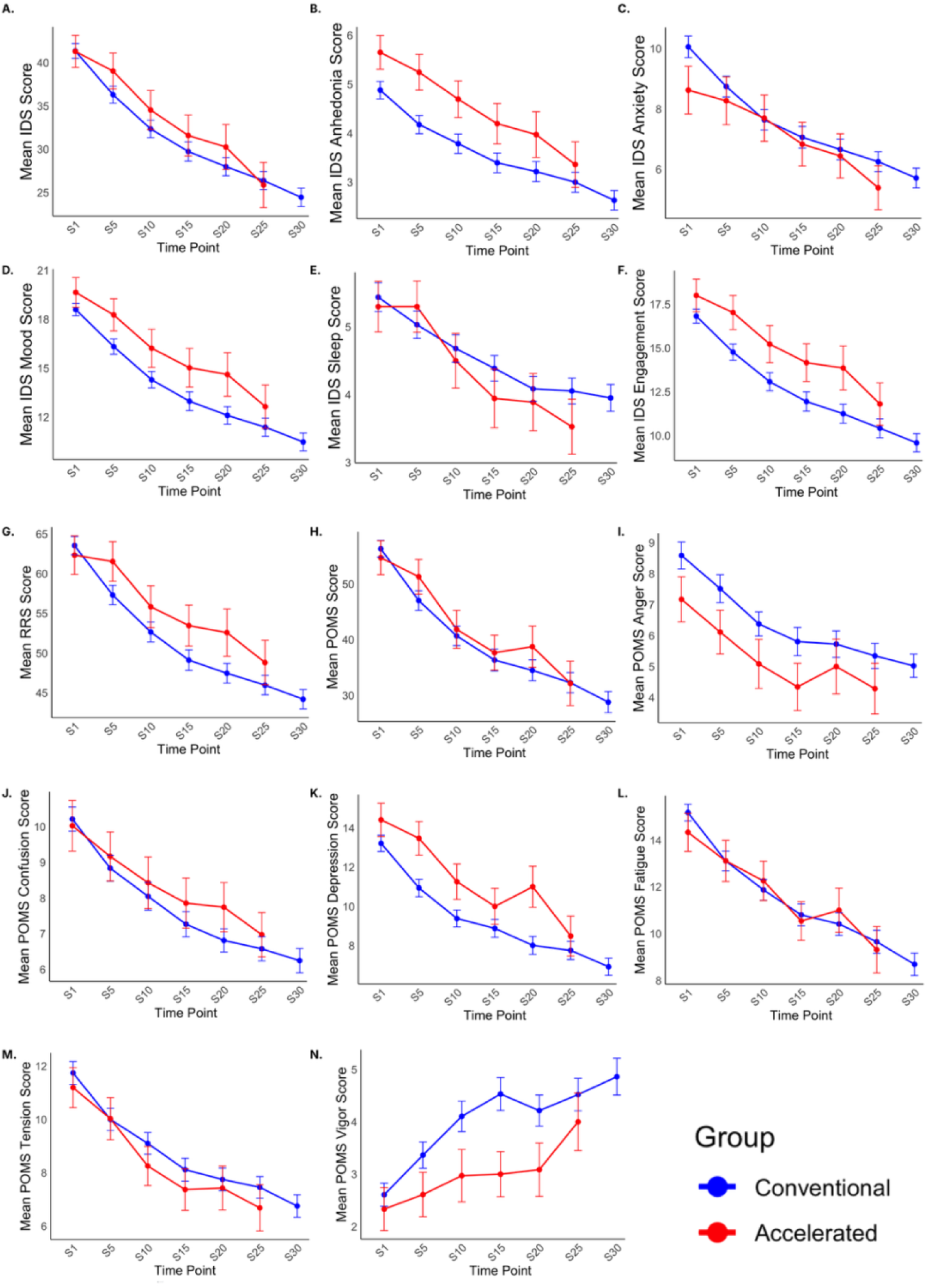
Trajectories of the IDS (Figure 1A), IDS subscales (Figures 1B-F), RRS (figure 1G), POMS (figure 1H), and POMS subscales (figures 1I-N), from before treatment (S1) to after treatment (S25 for accelerated patients and S30 for conventional patients. Accelerated patients are shown in red lines and conventional patients are shown in blue lines. Both conventional and accelerated rTMS protocols improved all 14 depression symptom domains. Note: S = “Session”.

Mixed-effects linear models showed that across all 14 depression symptom domains, there were significant main effects of timepoint (all *p*-values < .05, Bonferroni corrected), suggesting that both protocols led to improvements in depression symptom subscales from pre-to post-rTMS (Figure 2). There were no significant main effects of treatment type (all *p*-values > .05), indicating there were no significant differences between conventional and accelerated 5x5 rTMS when averaging across timepoints. Additionally, there were no statistically significant interaction terms (all *p*-values > .05), suggesting that group differences in changes across the IDS, IDS subscales, RRS, POMS, and POMS subscales were not detected. See Table 2 for a summary of these models.

**Figure 2.**
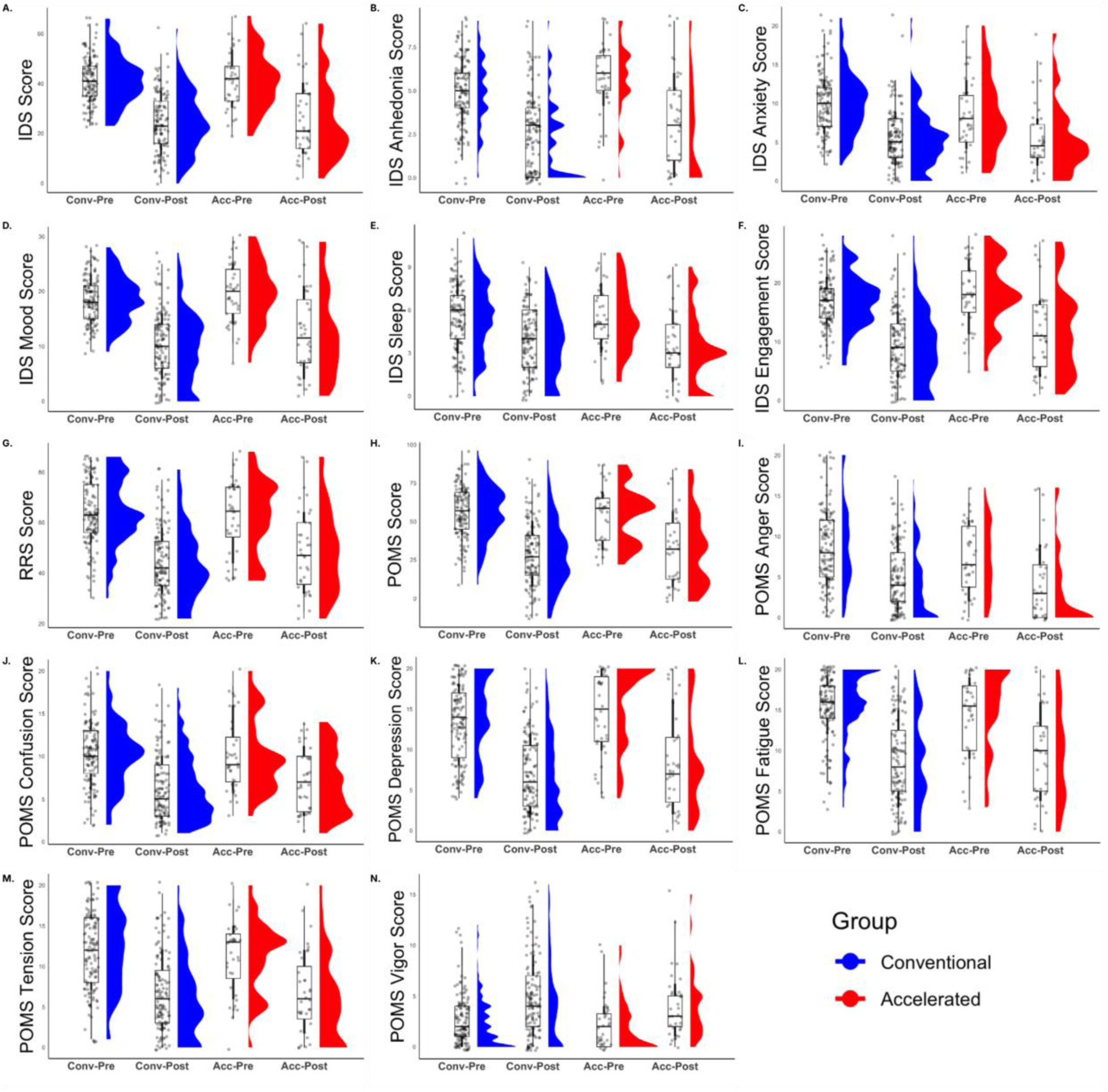
Pre- and post-rTMS scores for the IDS (Figure 2A), IDS subscales (Figures 2B-F), RRS (figure 2G), POMS (figure 2H), and POMS subscales (figures 2I-N). In each figure, the two box-and-whisker plots on the left correspond to the conventional group and the two box-and-whisker plots on the right correspond to the accelerated group. Conv-Pre = conventional group, pre-rTMS score; Conv-Post = conventional group, post-rTMS score (session 30); Acc-Pre = accelerated group, pre-rTMS score; Acc-Post = accelerated group, post-rTMS score (session 25). Conventional patients are shown in blue and accelerated patients are shown in red.

**Table 2.**
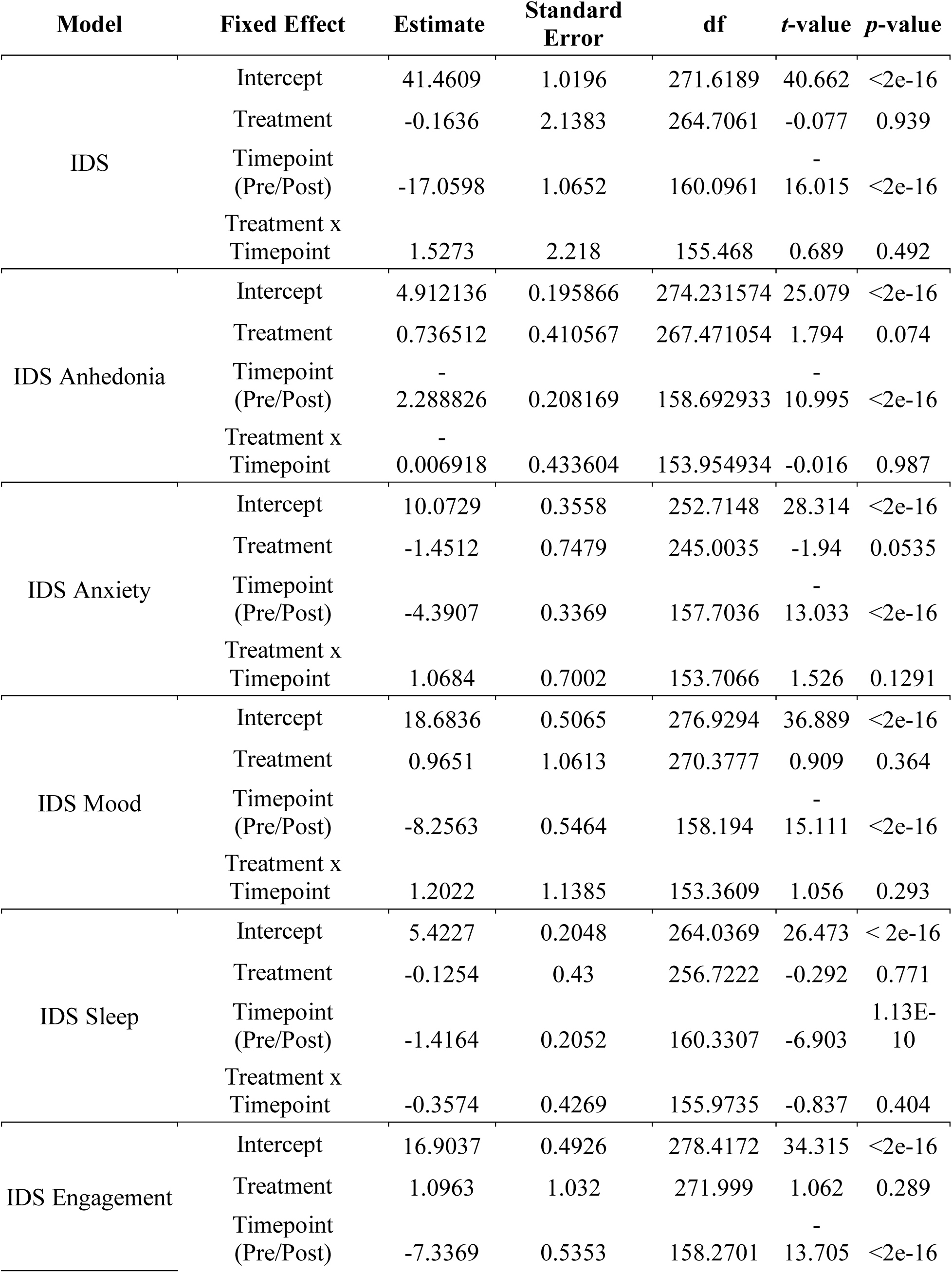
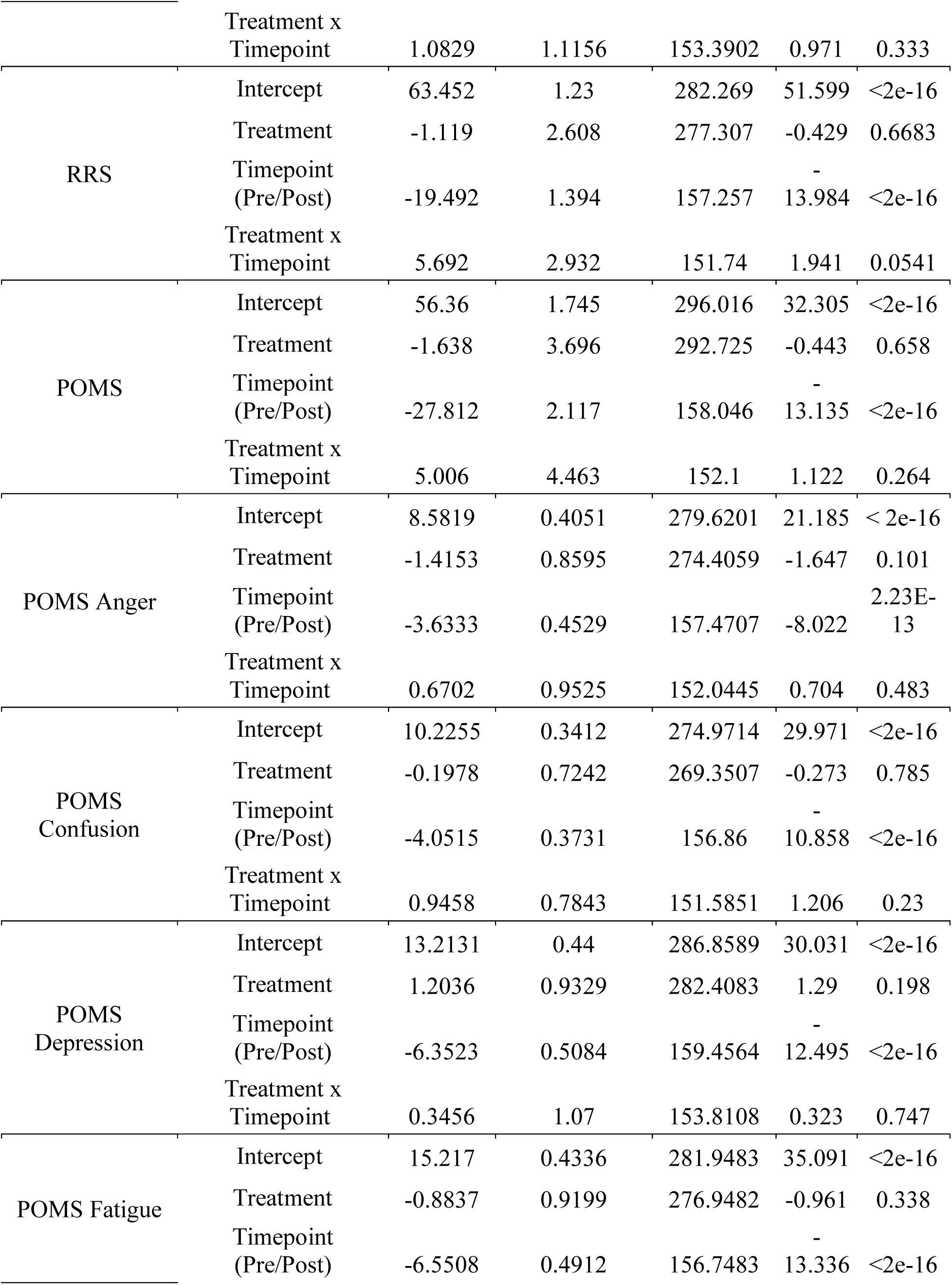
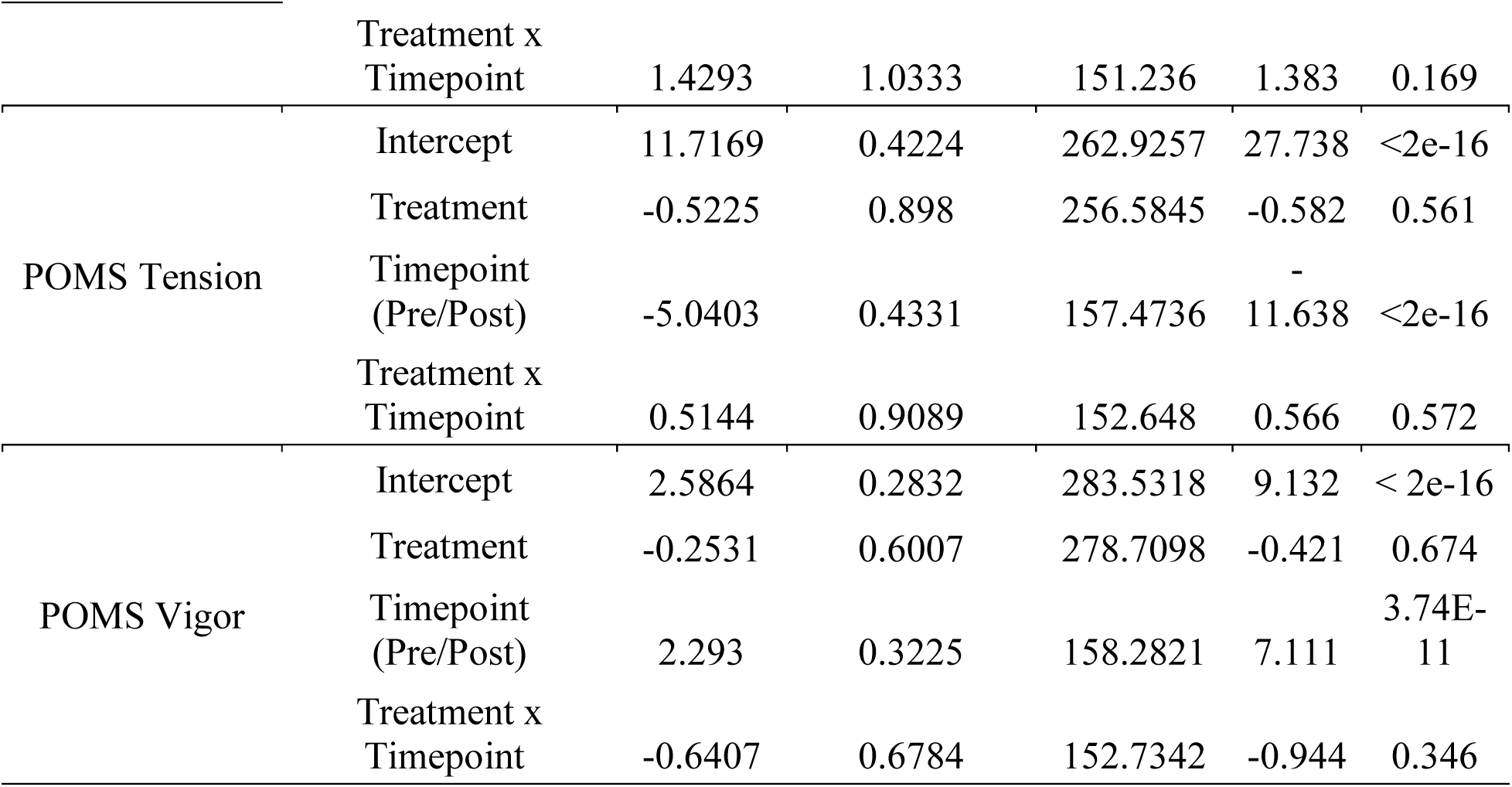
Mixed-effects linear models predicting the outcome variables (Model column) with Treatment (conventional and accelerated), Timepoint (Pre = S1 and Post = S25 for accelerated patients and S30 for conventional patients), and Treatment x Timepoint (interaction term) as fixed effects, and participants as random effects. Note: IDS and RRS represent the total scores for those subscales.

Exploratory models testing longitudinal changes in depression symptom domains showed that for the IDS overall score and subscales (anhedonia, anxiety, mood, sleep, and engagement), RRS, POMS overall score and subscales (anger, confusion, depression, fatigue, tension, and vigor), there were numerical differences across timepoints (all *p*-values < .05, uncorrected), indicating that symptom severity differed across longitudinal assessment points during rTMS treatment, averaged across the two treatment groups. For the IDS anxiety subscale, there was a main effect of treatment (*p* = 0.047, uncorrected), suggesting that anxiety symptoms were greater in the conventional group compared to the accelerated group. The following treatment x timepoint interactions were observed (uncorrected *p*-values): IDS anxiety subscale scores following treatments 10 (*p* = 0.017), 15 (*p* = 0.048), and 20 (*p* = 0.048); RRS scores following treatments 5 (*p* = 0.012), 15 (*p* = 0.021), and 20 (*p* = 0.007); POMS scores following treatment 5 (*p* = 0.048); POMS depression subscale scores following treatment 20 (*p* = 0.046); POMS vigor subscale scores following treatments 10 (*p* = 0.041), 15 (*p* = 0.005), and 20 (*p* = 0.046). For these treatment x timepoint interactions, conventional rTMS led to greater longitudinal improvements (see Figure 1). These analyses were exploratory and computed without a multiple comparison correction. See Table 3 for a summary of these models.

**Table 3.**
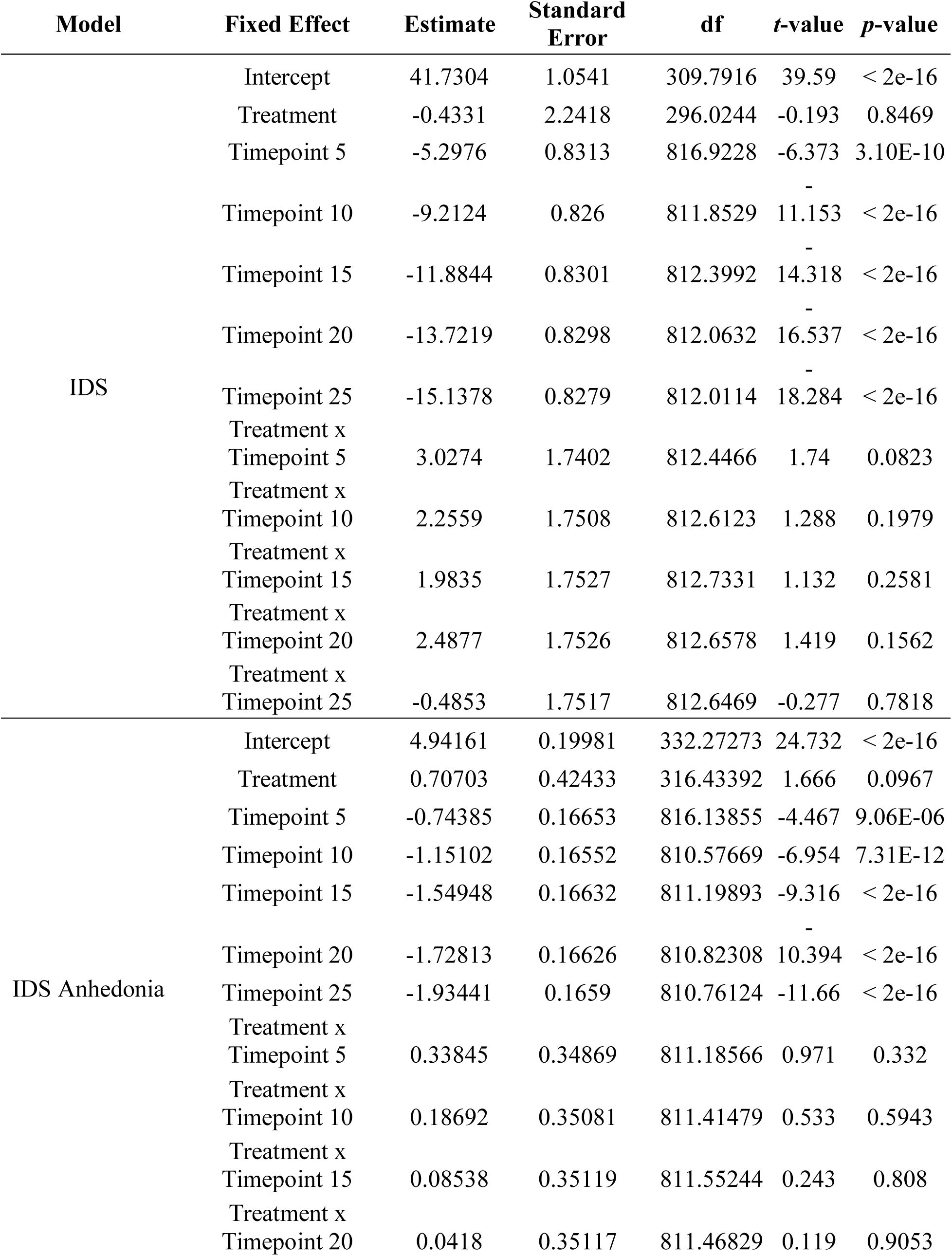
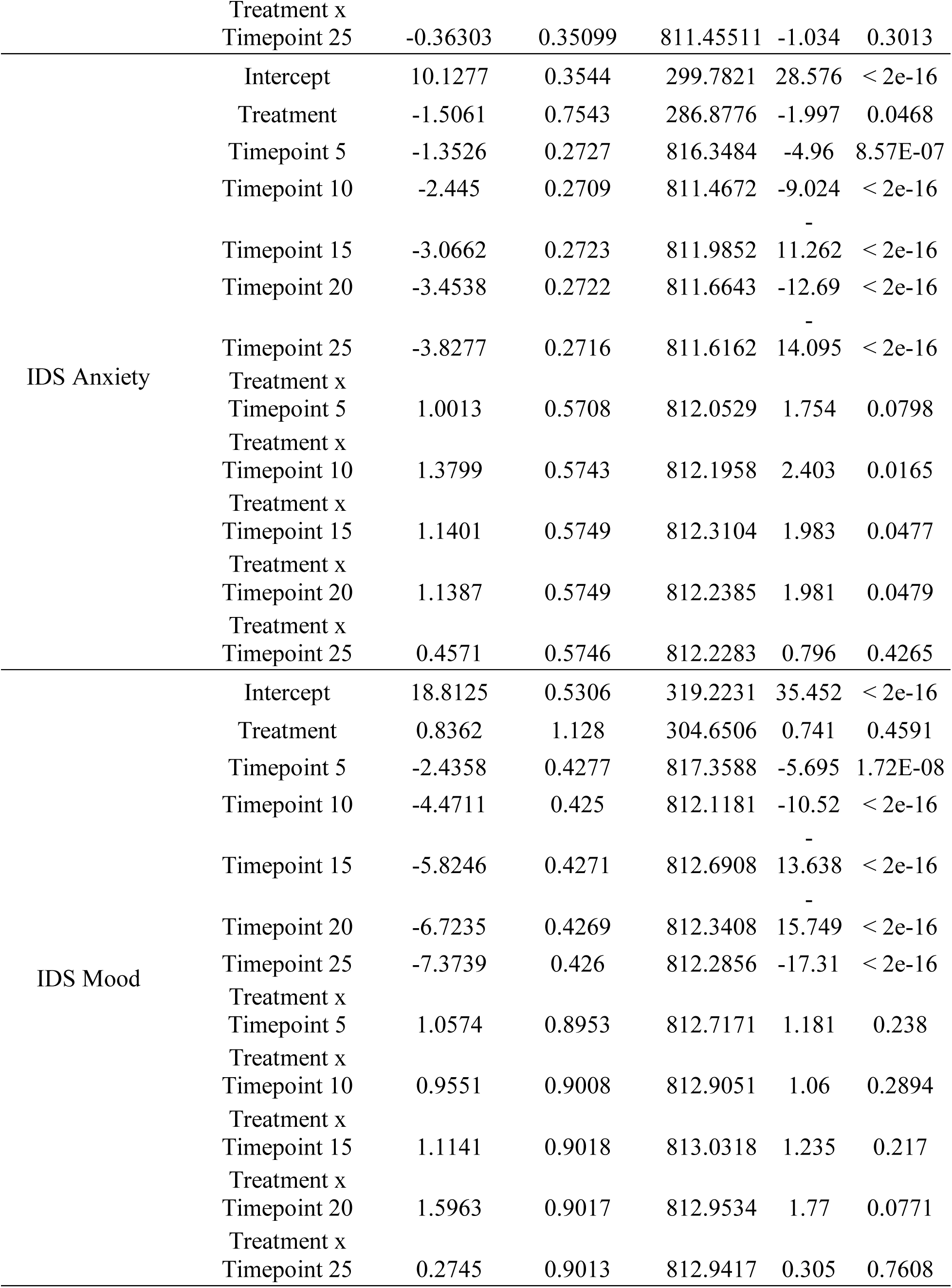
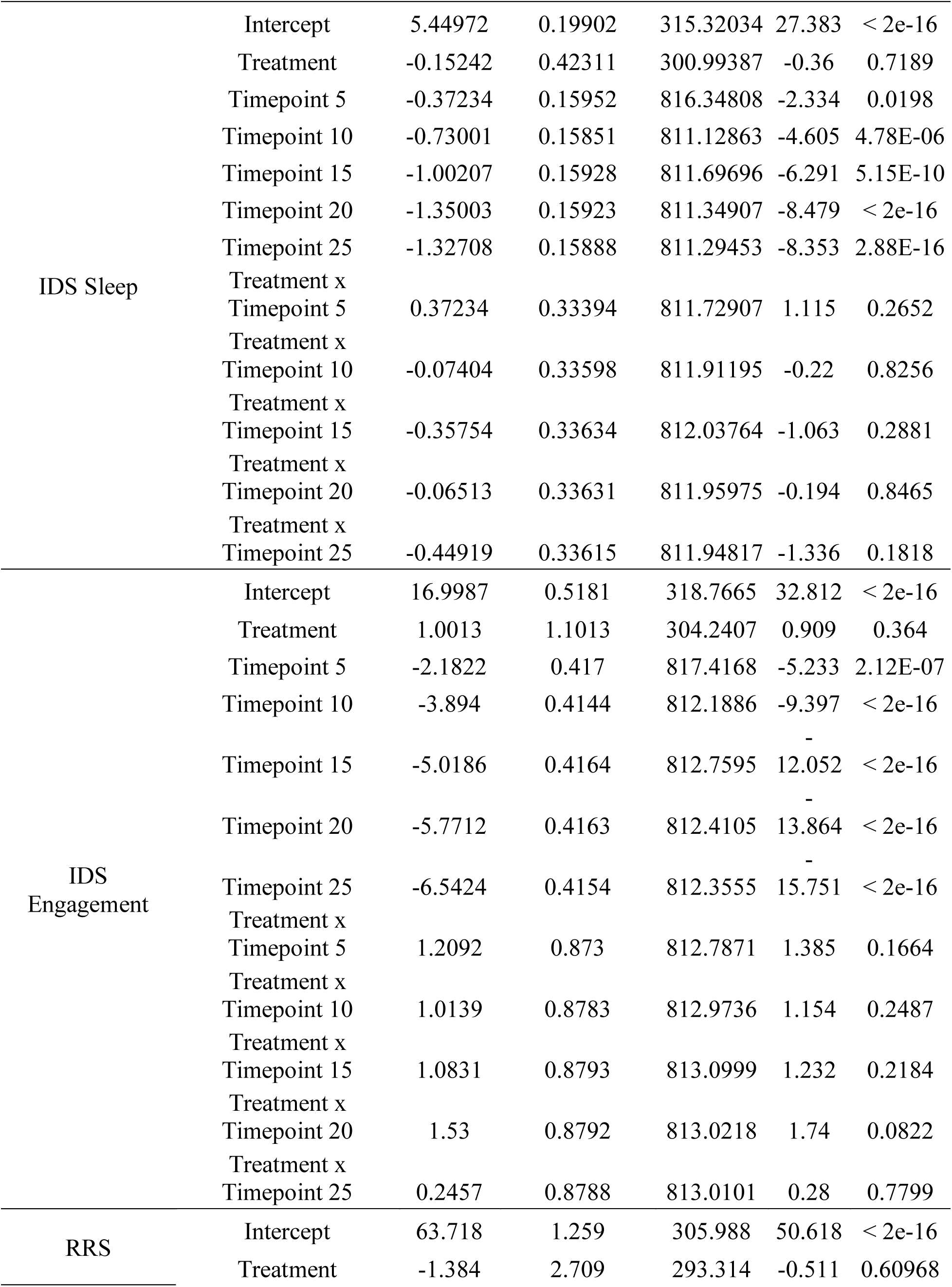
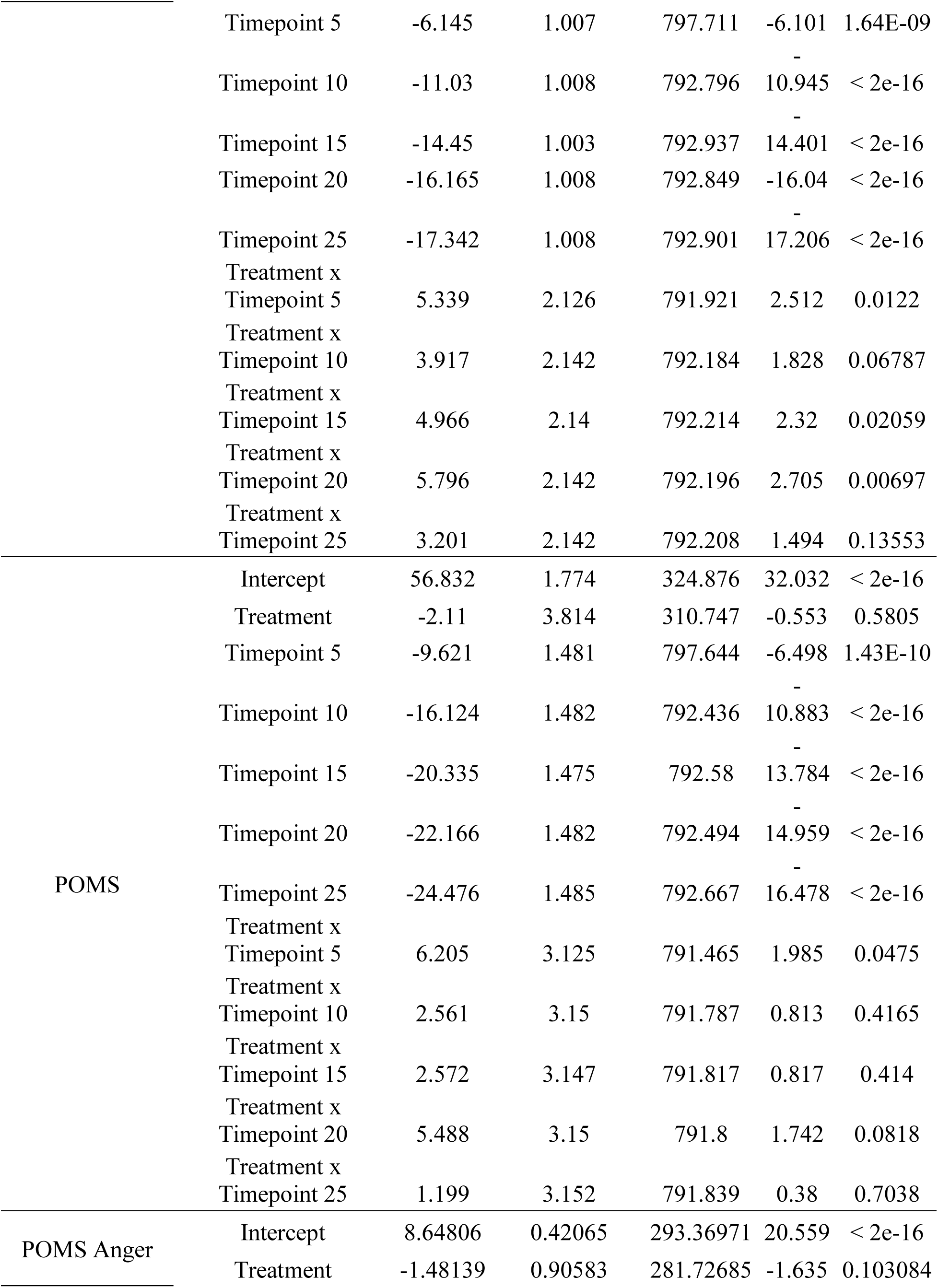
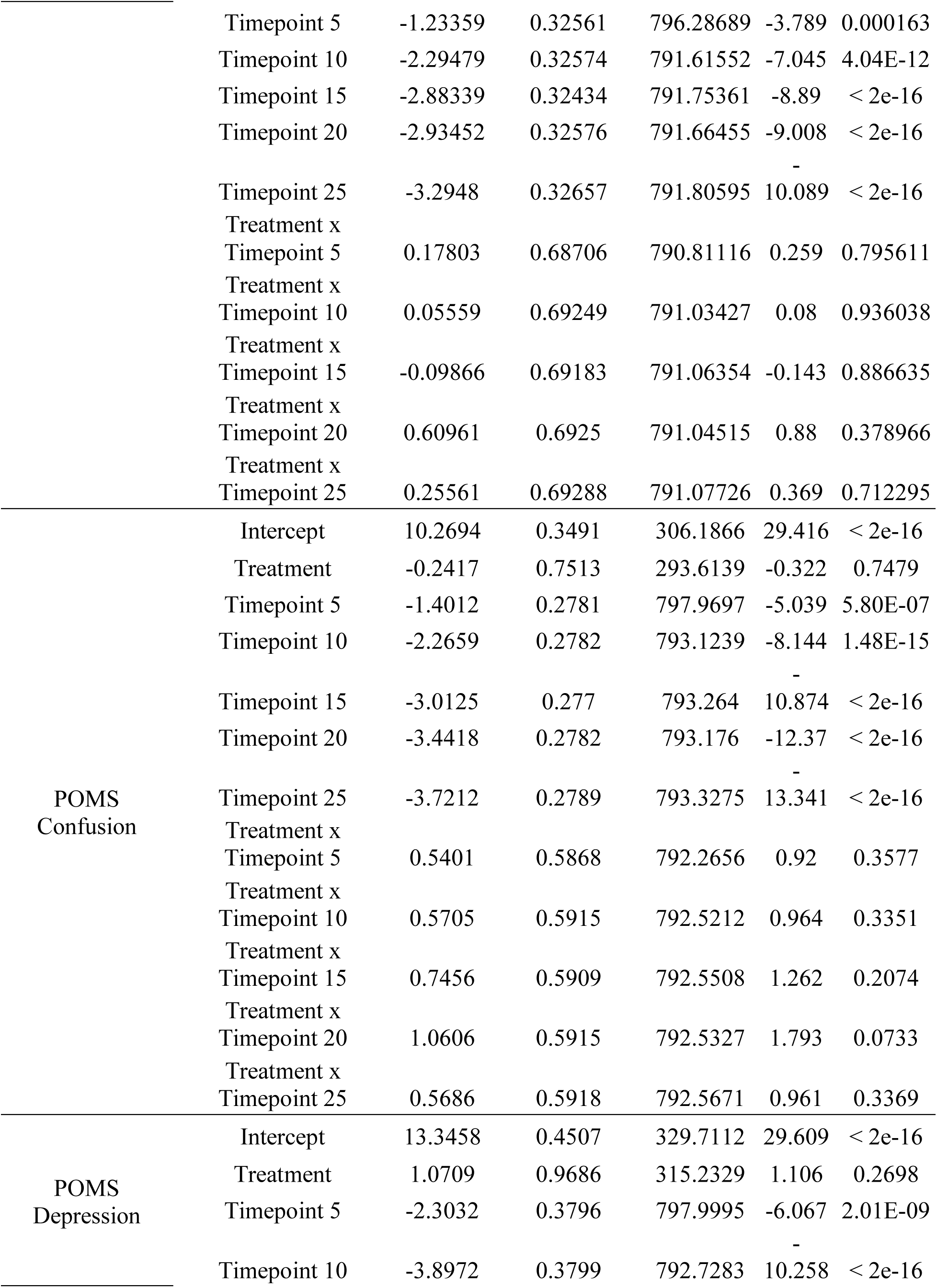
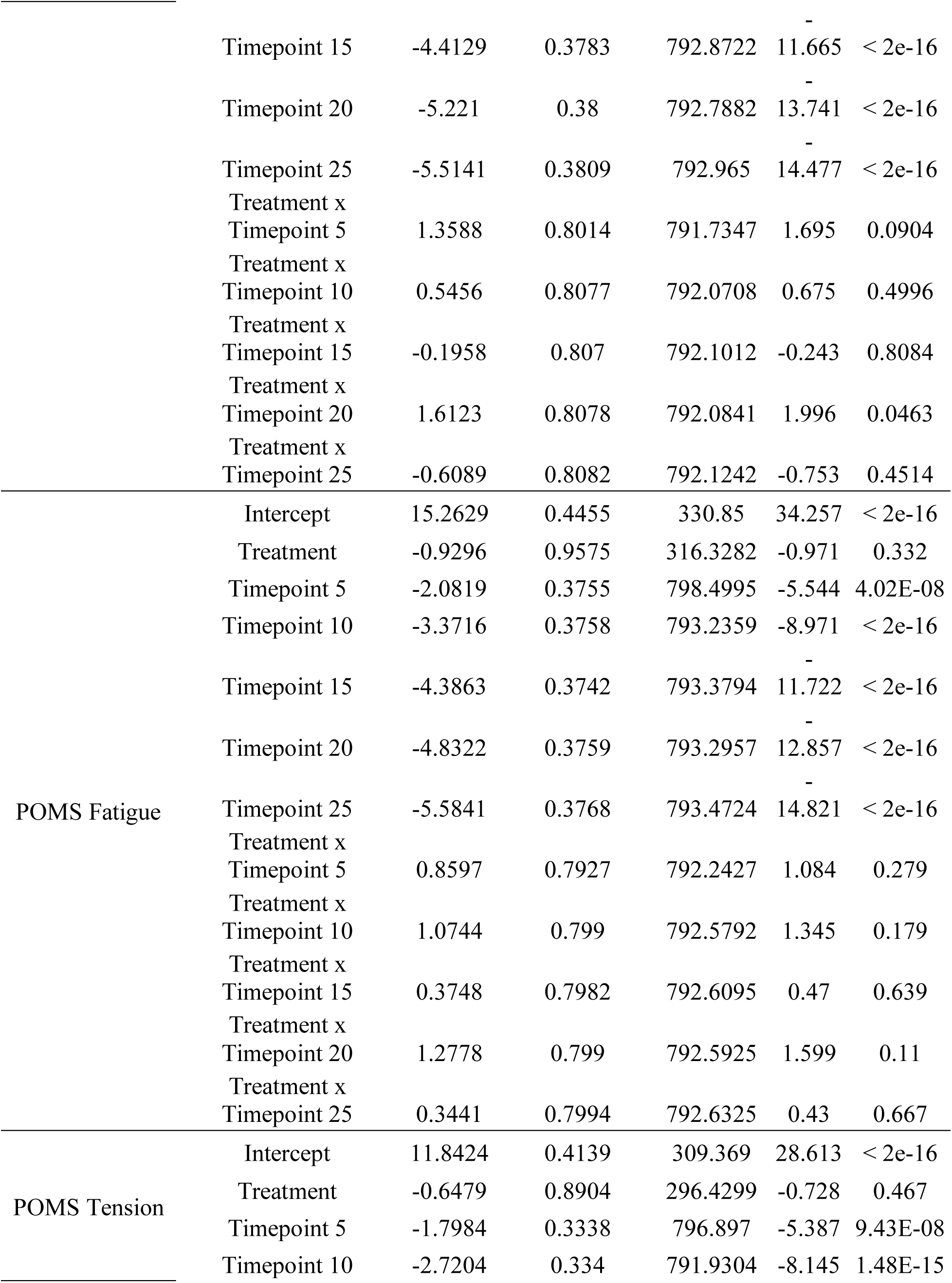
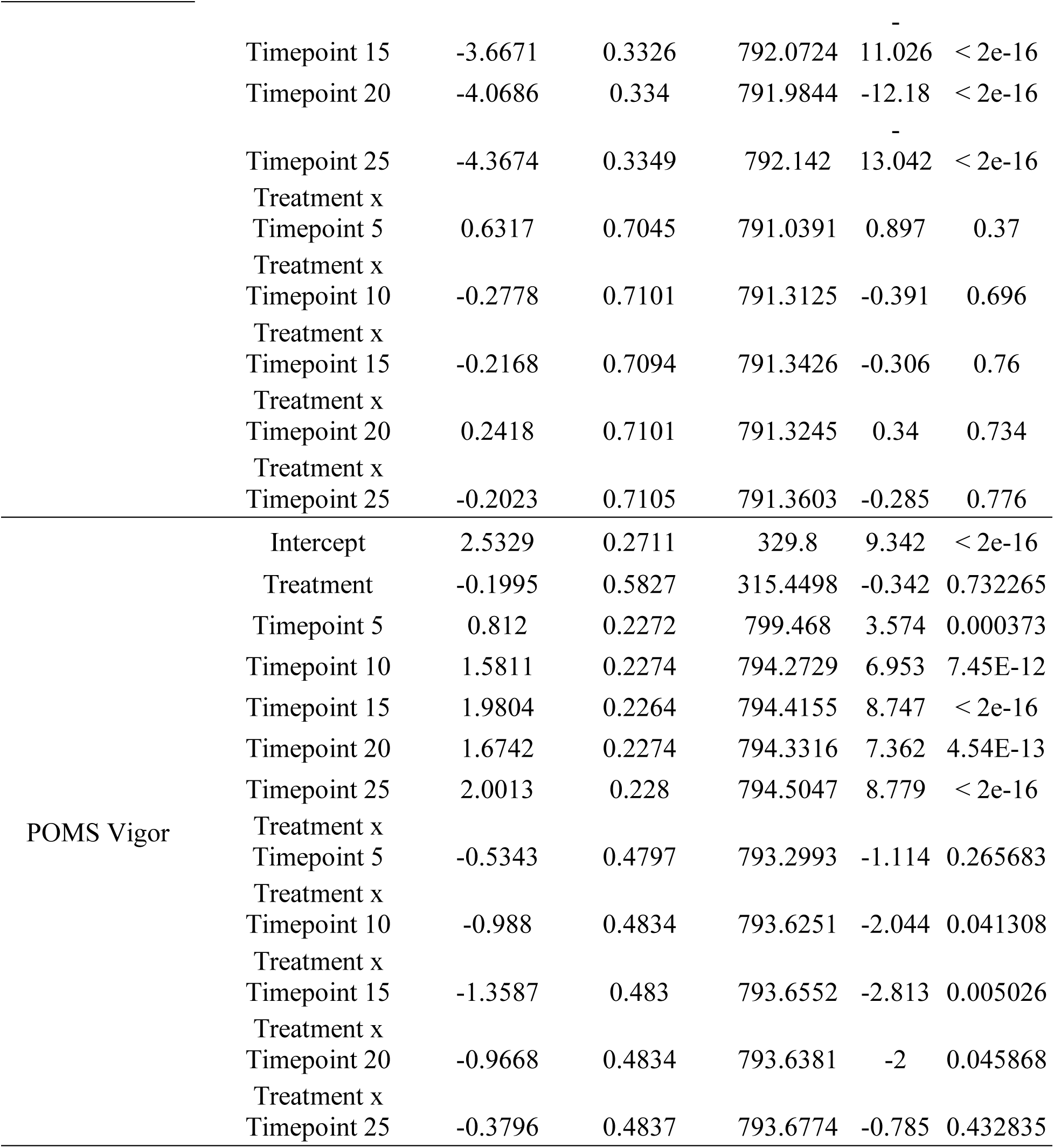
Mixed-effects linear models predicting the outcome variables (Model column) with Treatment (conventional and accelerated), Timepoint (session 5 to session 25), and Treatment x Timepoint (interaction term) as fixed effects, and participants as random effects. The reference categories were conventional rTMS for treatment and timepoint 0 (baseline) for timepoint.

## Discussion

The goal of this study was to compare the efficacy of conventional and accelerated 5x5 rTMS in alleviating distinct domains of depression symptoms. The primary analyses indicated that both rTMS protocols led to significant clinical improvement in depression symptom subscales that measured anhedonia, anxiety, mood, sleep, life engagement, rumination, anger, confusion, depression, fatigue, tension, and vigor. There were no statistically significant interaction terms for the primary analyses, suggesting that conventional and accelerated 5x5 rTMS had comparable efficacy in ameliorating depression symptom domains measured by the IDS, RRS, and POMS. This is consistent with prior work that has found similar efficacy between conventional and accelerated 5x5 rTMS in reducing overall depression levels (Apostol et al., 2026), as well as between conventional rTMS and other accelerated rTMS protocols (Caulfield et al., 2022).

The current results extended previous work by demonstrating comparable efficacy between the two protocols on a wide range of depression symptom domains, rather than overall sum depression scores. Notably, improvements in depression symptom domains occurred in five days in the accelerated 5x5 treatment, in comparison to 6 weeks for patients who received the conventional protocol. This supports the use of accelerated 5x5 rTMS as a faster acting intervention without diminished efficacy in alleviating depression symptom domains, suggesting that it is an appropriate choice for patients with varying symptom profiles.

Although there were no significant differences observed in the primary analyses, exploratory analyses that modeled symptom changes throughout a treatment course suggested that there may be clinically meaningful timepoint-by-treatment interactions. The exploratory results suggested that differences in symptom improvement during treatment may be observed for anxiety symptoms (IDS anxiety subscale) at treatments 10, 15, and 20, rumination (RRS) at treatments 5, 15, and 20, mood symptoms (POMS) at treatment 5, depression (POMS depression subscale) at treatment 20, and feelings of vigor (POMS vigor subscale) at treatments 10, 15, and 20. Given the exploratory nature of these models and the large number of tests without multiple comparison correction, these effects should be interpreted conservatively and are considered hypothesis-generating rather than hypothesis-confirming. Future randomized controlled trials could use these results to generate hypotheses about treatment stratification to conventional versus accelerated rTMS protocols based on symptom presentation.

One potential explanation for the comparable effects observed between conventional and accelerated rTMS on depression symptom domains is the shared stimulation target. Both groups received stimulation to the DLPFC, which has myriad functions spanning executive function, inhibitory control, planning, decision-making, and goal-directed cognition (Funahashi, 2022). Functional neuroimaging work has demonstrated that the DLPFC is implicated in various cognitive and psychological processes including anger (Pawliczek et al., 2013), anhedonia (Kazemi et al., 2025), sleep (Bosch et al., 2013), and anxiety (Balderston et al., 2017), which may explain why the stimulation of the DLPFC can help alleviate multiple depression symptom domains. It is possible that stimulation of other treatment targets would have led to differential reductions in the domains of symptoms measured here. For instance, stimulation of the orbitofrontal cortex might have been more efficacious for patients with higher levels of anhedonia (Feffer et al., 2018).

## Limitations and Future Directions

This work is not without limitations. This study was not a randomized clinical trial and in the absence of prospective treatment assignment, it is possible that other clinical or demographic factors could have contributed to the results. Because of the measurement-based care paradigm used in the conventional treatment group, there may have been variability in the treatment parameters between subjects based on clinician judgment (Einstein et al., 2026; M. K. Leuchter et al., 2023; Slan et al., 2024; Tadayonnejad et al., 2023; Berman et al., 2024; Lee et al., 2020; Citrenbaum et al., 2023; M.K. Leuchter et al., 2024). Similarly, conventional patients received 30 rTMS sessions and accelerated patients received 25, which should increase caution when interpreting the results. For the accelerated 5x5 patients specifically, some received RF-rTMS after consenting to participate in another study, and others received piTBS, and there may have been systematic differences between these patients, although past work demonstrated that there were no significant differences in improvement between RF and piTBS accelerated patients (Apostol et al., 2026).

A multiple comparison correction was applied for the primary analyses but not for the exploratory analyses. As such, the exploratory analyses should be interpreted cautiously and as hypothesis-generating rather than hypothesis-confirming. Future randomized clinical trials are needed to confirm the differential effects of conventional and accelerated rTMS on depression symptom domains. Furthermore, the IDS, RRS, and POMS were self-report measures and subject to bias (Fukuda et al., 2021; Petrowski et al., 2021; Rush et al., 1996; Thase et al., 2023; Townshend & Hajhashemi, 2023; Trivedi et al., 2004; Wardenaar et al., 2010). Clinician-rated measures could be included in future work to address this limitation. Although the subscales captured a variety of depression symptoms experienced by patients with MDD, there was some overlap in the scale items included in some of the subscales, which is an important consideration when interpreting the results of this study.

Future investigations should consider alternative methods of characterizing depression symptom domains. For instance, unsupervised machine learning approaches can be used to identify MDD domains and could be incorporated in future work (Sharma et al., 2024). Other future studies should map the trajectories of depression symptom domains after the completion of rTMS; this will provide more information on the durability of conventional versus accelerated rTMS in alleviating specific depression symptoms. If future studies demonstrate that there are some symptoms that do not respond as well to a specific rTMS protocol, then it would warrant the exploration of other stimulation targets or protocols to provide more comprehensive care to patients with varying symptoms. Finally, future investigations into biomarkers that predict response to both conventional and accelerated rTMS protocols is an important future direction, and studies should focus on magnetic resonance imaging (Morriss et al., 2024; Oathes et al., 2023), EEG (A. F. Leuchter et al., 2021), or other physiological measures such as pupillary reactivity (Citrenbaum et al., 2023). Additionally, due to the vast rTMS parameter space, future empirical investigations should compare heuristic versus neuronavigated targeting (Fitzgerald et al., 2009; Hebel et al., 2021; Kinjo et al., 2024), different coil types (Zibman et al., 2021), and rTMS alone versus rTMS combined with medication or talk therapy (Zaidi et al., 2024), in relation to specific depression symptom domains.

## Conclusion

This study demonstrated that conventional and accelerated 5x5 rTMS have comparable efficacy in improving a variety of depression symptom domains. This work addressed concerns in the literature that depression sum scores do not fully encompass symptom variability between patients (Fried & Nesse, 2015a, 2015b; Marx et al., 2023). Future studies should implement similar approaches in modeling MDD symptom domains, which will help generate a more nuanced understanding of how different rTMS protocols may benefit patients with different symptom profiles. Decisions on whether to pick one protocol versus another will depend on a number of factors including patient preference, clinician expertise, staff training, and equipment availability. Together, these findings suggest that conventional and accelerated rTMS have comparable efficacy in reducing the severity of a wide variety of depression symptoms. Future investigations should extend this work and further explore how patients with different symptom profiles respond to different rTMS protocols. This is a step towards advancing precision psychiatry and rTMS treatments for MDD.

## Data Availability

Data are not available for sharing due to patient confidentially.

